# Is it time for the neurologist to use Large Language Models in everyday practice?

**DOI:** 10.1101/2025.01.23.25320945

**Authors:** N. V. Maiorana, S. Marceglia, M. Treddenti, M. Tosi, M. Guidetti, M.F. Creta, T. Bocci, S. Oliveri, F. Martinelli-Boneschi, A. Priori

## Abstract

Large Language Models (LLMs) such as ChatGPT and Gemini are gaining momentum in healthcare for their diagnostic potential. However, their real-world applicability in specialized medical fields like neurology remains inadequately explored. The possibility to use these tools in everyday diagnostic practice relies on the evaluation of their ability to serve as support for the clinician in assessing the patient, understanding the possible diagnosis and design the diagnostic pathway. To this end, in this study we (1) examined the available literature on the evaluation of LLMs in neurology diagnosis in order to understand whether the methodologies applied were adequate to translate the use of LLMs in everyday practice, and (2) designed and performed an experiment to evaluate the diagnostic accuracy and clinical recommendations of ChatGPT-3.5 and Gemini compared to neurologists using real-world clinical cases presented following the everyday diagnostic practice. In the vast literature of LLMs application in neurology, only 24 studies reported experiences using LLMs in clinical neurology. The experiments reported showed a heterogeneous scenario of prompt engineering and input formats. At present, while responses using structured prompts were well documented, there is a lack of studies using real-world clinical scenarios, and everyday workflows and practice. We therefore conducted a real-world experiment using a cohort of 28 anonymized patient records from the neurology department of the ASST Santi Paolo e Carlo Hospital (Milan, Italy). Cases were presented to ChatGPT-3.5 and Gemini replicating the typical clinical workflows. Diagnostic accuracy and appropriateness of recommended diagnostic tests were assessed against discharge diagnoses and neurologists’ performance.

Neurologists achieved a diagnostic accuracy of 75%, outperforming ChatGPT-3.5 (54%) and Gemini (46%). Both LLMs exhibited difficulties in nuanced clinical reasoning and over-prescribed diagnostic tests in 17–25% of cases. Despite their ability to generate structured recommendations, they struggled with complex or ambiguous presentations, requiring additional prompts in some cases. We can therefore conclude that LLMs have potential as supportive tools in neurology but they currently lack the depth required for nuanced clinical decision-making. The findings emphasize the need for further refinement of LLMs and the development of evaluation methodologies that reflect the complexities of real-world neurology practice.

## Introduction

In recent years, artificial intelligence applications have revolutionized the technological landscape with potential consequences also in the medical field ^1^. Among them, large language models (LLM) and Generative Pre-trained Transformer (GPT) implementations have significantly enhanced the possibilities for a fluid and natural human robot interaction ^1^.

LLM systems are designed to understand and generate human-like text ^2^. They are trained on massive datasets, using deep learning techniques to predict the next word or phrase in a sequence ^2^, enabling them to perform tasks like summarization, translation, and conversation ^3^. GPT is a specific implementation of a LLM that uses an architecture called Transformer. This specific architecture allows to create LLMs able to handle large sequences of text efficiently by focusing on the relationships between words across a sentence, making them able to maintain context and generate responses that are coherent and human-like ^4^. A GPT model is initially trained on large-scale text datasets before being fine-tuned for specific tasks ^5^.

LLMs now readily available for everyday use, as Chat-GPT made by OpenAI and Gemini made by Google, are based on LLM-GPT technology. These advanced LLMs enable users to easily interact enhancing accessibility for both personal and professional purposes.

These generative language models, promise significant potential in assisting healthcare professionals, especially in diagnosis, medical treatments ^6^ and medical education ^7^. Physicians might receive rapid support in analyzing clinical data, interpreting results, and formulating hypotheses ^8^.

However, the adoption of LLMs such as Chat-GPT and Gemini also raises ethical considerations and potential risks ^9^.

As a first point, these models are trained on extensive but generalist datasets which may limit their performance in specialized domains ^10^.

The quality, origin, and type of training data may generate an inherent potential for bias, amplified in specialized fields like medicine that demand profound knowledge, understanding and critical thinking ^11^. To date there is a lack of control and transparency on the data used to train the models^12^ thus influencing the accuracy and reliability of the outputs ^13^.

Another point regards the excessive reliance on these tools that could lead to reduced human oversight and critical thinking which are of fundamental importance for medical decision-making ^14^. Additionally, concern resides in the risk that patients with direct access to these models may misinterpret symptoms or engage in self-diagnosis, potentially leading to negative health consequences ^15^. Responsible usage, therefore, must be promoted among healthcare professionals and patients, emphasizing a balanced integration between ai powered LLMs and humans agents ^13^. On the other hand recent studies highlighted the increasing role of LLM-GPTs, in the field of neurology, showing promising applications and fast progression across diagnostic and evaluative tasks ^16^. For instance, ChatGPT-4 was evaluated on neurology board-style exams, where it exceeded the performance of ChatGPT-3.5 and surpassed the human average, showing strength in both lower-order and higher-order reasoning tasks, suggesting potential uses in neurology education and diagnostic support ^17^.

ChatGPT diagnostic accuracy was also assessed in neuro-ophthalmology, finding that ChatGPT-4 reached an accuracy close to human expert performance ^18^. In neuro-oncology, ChatGPT-4 achieved good levels of accuracy in both diagnostic process and treatment planning, with high ratings from neurosurgeons, it was noted that further refinement is needed for clinical use ^19^. Lee et al. ^20^ explored Chat-GPT-4’s capability in lesion localization for stroke patients, reporting high precision and specificity, indicating its potential as a supplementary tool for neuroanatomical localization. Chat-GPT-4 was also examined as a neuro-score calculator for Glasgow Coma Scale, showing decreased accuracy and a major frequency of hallucination (e.g., suggesting nonexistent medical treatments) with increased complexity in input phrasing, underscoring limitations when interpreting complex clinical information ^21^.

Chat-GPT has proven beneficial in triage, producing diagnostic outcomes similar to those of healthcare professionals ^22^. These findings highlight the potential of LLMs to assist in clinical settings ^22^.

However, while LLMs have demonstrated proficiency in various neurological fields when assessed on well-known clinical cases depicted by educational vignettes or single cases reported in the literature ^18,19^, there is a lack of data examining their performances on real world cases from daily clinical practice of a neurology department: much of the existing literature on clinical decision-making tools and physician-assistive technologies relies heavily on carefully curated scenarios and inputs designed explicitly for research purposes. While these studies have undoubtedly contributed to our understanding of the potential capabilities of these systems, they often fail to capture the chaotic unpredictability of real-world clinical environments. The carefully crafted prompts and sanitized datasets typically used bear little resemblance to the high-pressure, overcrowded clinics where overworked physicians must make rapid decisions under suboptimal conditions. This discrepancy raises a critical question: How do these tools perform when faced with the unstructured, fragmented, and often overwhelming realities of clinical practice? Specifically, what happens when such systems are employed by a “lazy” doctor—one who, burdened by fatigue, systemic inefficiencies, and competing priorities, may be less meticulous in their interactions with these technologies?

Therefore, aiming to assess diagnostic accuracy and reliability of LLMs in real-world applications, in this study, (1) we examined the available literature on the evaluation of LLMs as diagnostic support in neurology, as already done in other fields, such as cardiology and ophthalmology ^23,24^, and (2) tested two of the most common, easy-to-reach, LLMs (Gemini made by Google and ChatGPT-3.5 made by Open AI) against neurologists in assessing real cases, in order to verify the competence of both LLMs in formulating relevant diagnostic hypotheses in the neurological field and recommending appropriate diagnostic tests. The underlying rationale is to break away from the idealized constructs of current research by simulating a realistic scenario: an overcrowded clinical world where physicians operate under relentless pressure. This simulation is not about testing how well systems function under perfect conditions; it is about understanding how these tools integrate into imperfect realities. By doing so, we hope to uncover critical insights into their usability, reliability, and potential pitfalls when deployed in the environments they are meant to serve.

### How is Large Language Models applicability in everyday neurology practice evaluated?

When evaluating LLMs in the field of neurology, it is crucial to consider that LLMs can be guided using different types of prompts, such as zero-shot prompts, which provide no prior task-specific guides, and few-shot prompts, including examples to help the model understand the request^25^. Structured prompts are highly detailed, specifying the context, the role of the model, and the desired output format, while iterative prompts involve a collaborative refinement process between the user and the model to optimize clarity and effectiveness^25^.

The format of input questions or text passed to the model—whether open-ended or multiple-choice—also significantly impacts the accuracy of LLM responses ^26^. Open-ended questions allow for a wide range of responses, which can lead to variability in accuracy depending on how well the prompt guides the model. Multiple-choice formats, however, constrain the model to select from predefined options, potentially improving accuracy by limiting the response scope^26^. Given these nuances, an assessment of the literature focusing on prompt types and input formats is crucial to establish methodologies for evaluating LLMs in neurology, ensuring that their integration into healthcare is both effective and ethically sound.

We therefore performed a literature search on PubMed (on December 1^st^, 2024), using the following search string: ((chatgpt) OR (chat-gpt) OR (gpt)) OR (gemini) AND neurology. The search was restricted to studies published between January 1^st^ 2019 to December 1^st^ 2024 (date of the search). Studies were included in the review if they utilized LLMs for diagnostic purposes in neurology or where their knowledge was assessed via neurology related questions. Exclusion criteria encompassed studies in formats such as opinion papers, reviews, meta-analyses, and technical papers, and non-peer reviewed paper. Furthermore, studies lacking a clear focus on neurology or those that did not evaluate LLMs in contexts relevant to the field were excluded. Finally, papers without sufficient methodological transparency to allow for the extraction of key variables, including prompt type, input type, and model specifications, were not considered in the review. In this review, zero-shot and few-shot prompts have been considered examples of *soft prompting*, as they rely on minimal contextual engineering and mimic natural communication. Conversely, structured and iterative prompts are categorized as *hard prompting*, given their high degree of detail and systematic design to guide the model’s responses effectively. Furthermore, studies in which questions were presented in multiple-choice format were considered to be hard prompted, as multiple-choice questions create a context that influences the interpretation of a specific request by the user. The types of cases used in the studies were recorded and categorized into real cases, simulated cases, and questions, based on the specific material provided to the LLMs. A total of 201 references were initially identified. Duplicate records were removed (n = 10), resulting in 191 unique references. Two reviewers (NVM and MG) independently screened the titles and abstracts, excluding opinion papers, reviews, meta-analyses, methodological studies, implementation-focused papers, and articles unrelated to neurology. The final sample consisted of 24 studies ^16,17,20,21,27–46^ (Table 1), which were included for data extraction. Three main case types were identified: standardized questions were used in 11 studies, real cases in 10 studies, and simulated cases in 3 studies. Prompt usage varied significantly across studies. Hard prompts were the most frequently employed, appearing in 18 studies. Studies involving multiple choices were considered as hard prompted due to the fact that multiple choices questions crate a context which can lead the LLMs a reference for the response. Hard prompts were used in 7 studies involving real cases and in 3 studies involving simulated cases. Soft prompts were used in 5 studies and only in 2 studies assessing models’ accuracy with real cases. The type of input provided to the LLMs varied across studies. Only three studies featured soft prompting, open ended input and real cases. Results showed a predominance of ChatGPT-4 and ChatGPT-3.5 as the models of choice across the studies. However, 12 out of 24 studies employed combinations of multiple LLMs to enable comparative analyses. Only two studies combined a soft approach, open ended question and real cases to compare the accuracy of at least two LLMs. The best performance was recorded by ChatGPT-4 in a study conducted by Abbas et al. (2024) ^39^, where structured prompts and multiple-choice inputs were used. ChatGPT-4 achieved 100% accuracy in diagnostic questions, outperforming ChatGPT-3.5, Claude, and Bard, highlighting its reliability in controlled and well-defined tasks. Conversely, the worst performance was observed in a study by Finelli (2024) ^28^, which employed open-ended prompts and real-case inputs. ChatGPT-4 failed to provide any correct diagnoses, while Glass AI achieved only one correct diagnosis. The analysis highlighted a methodological reliance on hard prompts, particularly in studies involving real cases and standardized questions. This preference underscored the utility of hard prompts in testing LLMs’ capacity to handle structured and complex clinical scenarios. However, the limited use of soft prompts, particularly with real cases, suggests an area requiring further exploration. Soft prompts could provide a more realistic simulation of everyday clinical interactions, better reflecting the variability and nuance inherent in neurology practice. A study incorporating soft prompts with real cases would significantly enhance the ecological validity of these evaluations. This methodological diversity is crucial for building a comprehensive understanding of LLM capabilities in neurology.

**Table 1.**
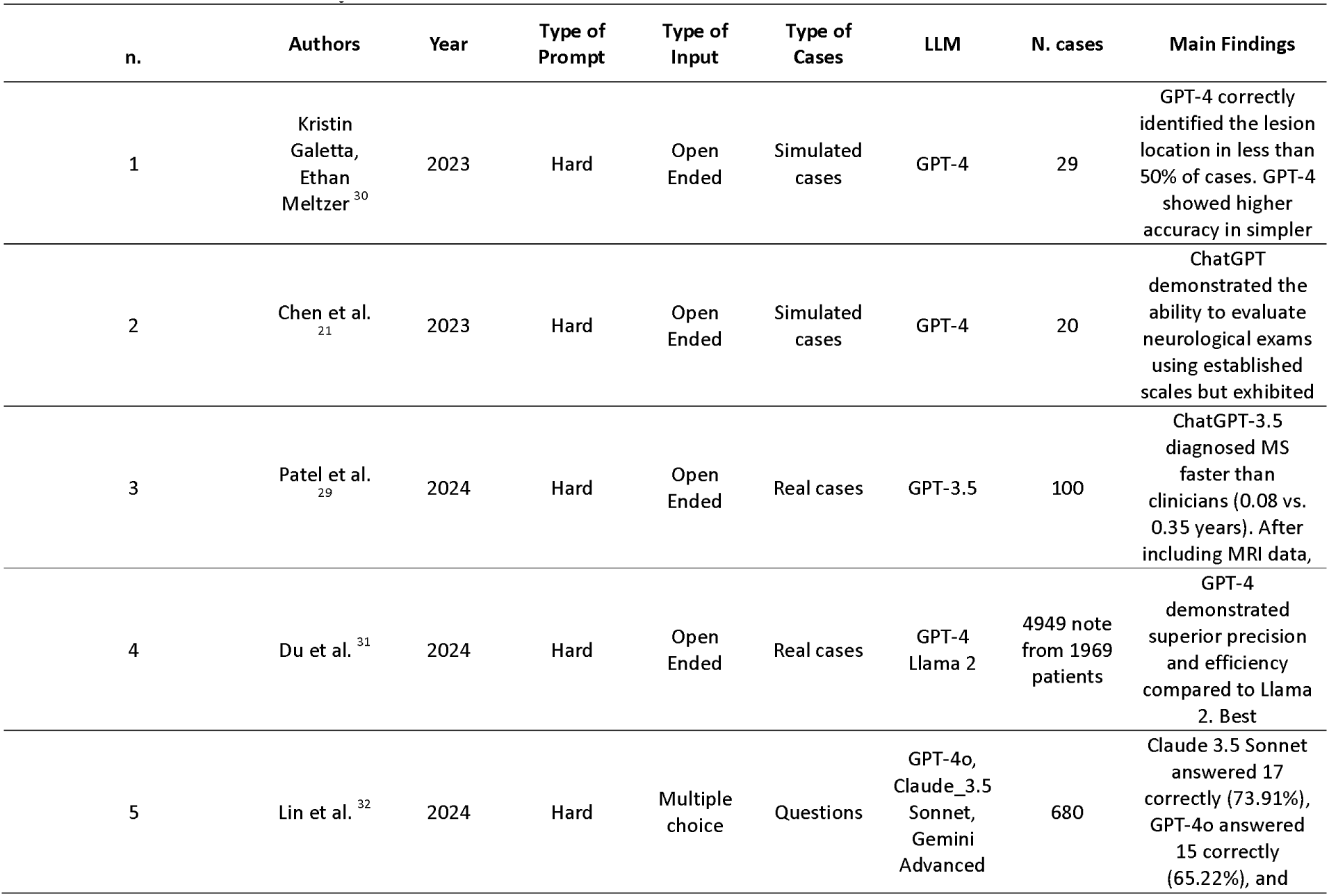
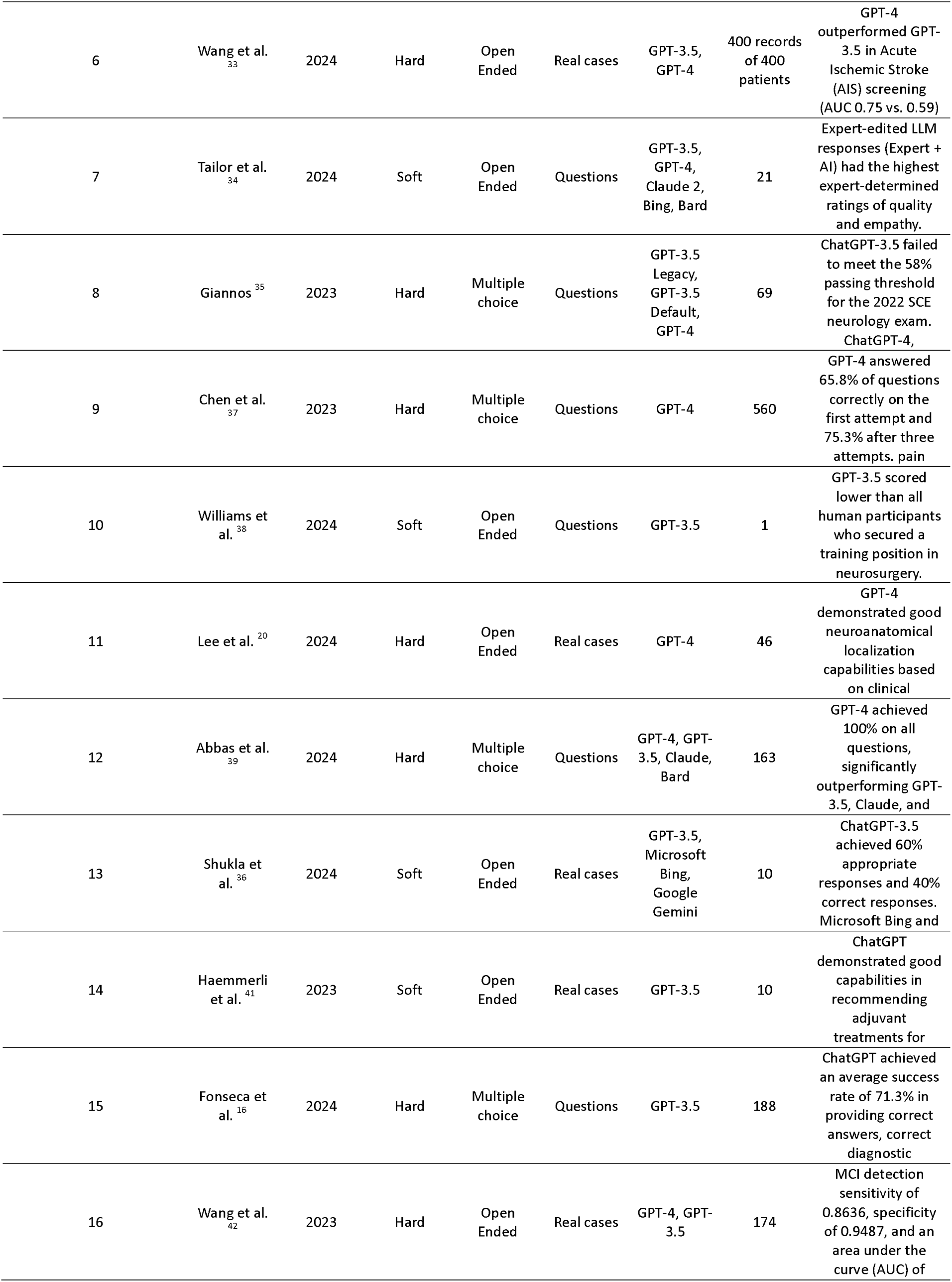
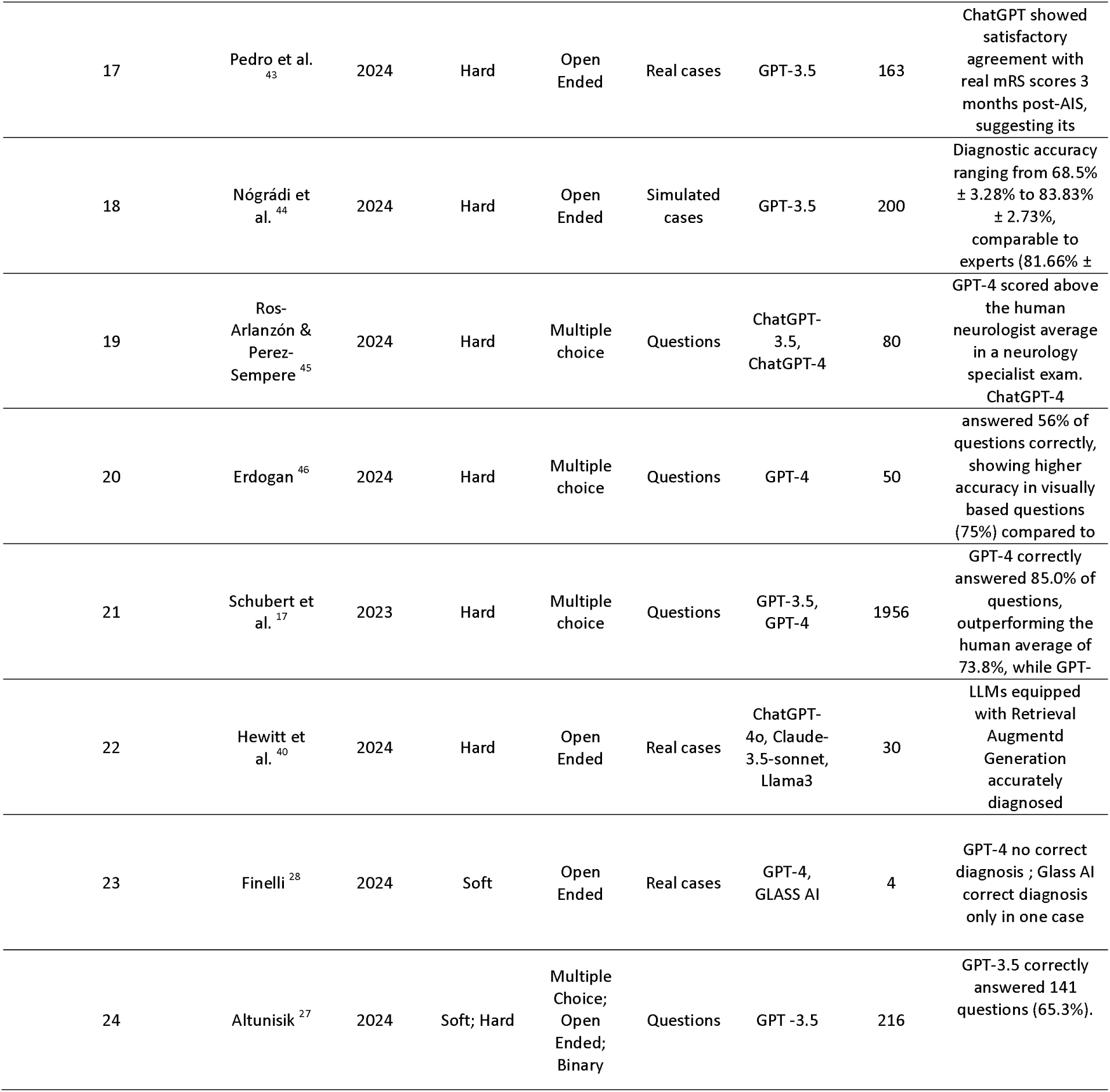
Extracted and analyzed studies.

### A real-world simulation scenario: the *“Lazy Doctor”* in an overcrowded clinical world

#### Methods

A total of 56 consecutive patients admitted to the University Hospital Santi Paolo e Carlo - Neurology department (Milan) between 1^st^ July and 31^th^ August 2023 were analyzed. Inclusion criteria were that cases should refer to patients who were admitted by a neurologist with a suspected diagnosis, cases should include a discharge letter containing the investigations conducted during the admission and a definitive diagnosis. Patients whose admission was purely therapeutic or observational, without a diagnostic purpose, were excluded. Additionally, patients with an already-known diagnosis were excluded.

The present study qualifies as a non-interventional observational study and does not constitute a clinical trial. The study has been approved by the IRB of the University of Milan with the approval number 123/24, All. 3 CE 10/12/24.

Patients provided consent for the processing of their personal data at the time of hospital admission under protocol ast_daz_502_ed00, in accordance with privacy regulations (D.Lgs. 101/2018, implementing EU GDPR 2016/679). This consent permits research on clinical data. The data have been anonymized prior to processing, ensuring that they cannot be traced back to any specific patient.

#### Simulation scenario

We retrieved the Electronic Health Records (EHRs) of the selected patients (pdf format) as they were filled in at the time of the patient’s admission. We simulated the process of the initial diagnosis considering the procedure depicted in Figure 1. The patient, coming from the ER, is admitted to the Neurology ward. The neurologist in charge of admitting the patient begins the examination by collecting: family history, history of present illness, past medical history, and reviews the diagnostic results performed in the ER. Additionally, the neurologist examines lab analysis results when available. For each selected patient, the clinical data available to the neurologist at the time of admission were collected. Not all EHRs were structured identically. Some patients had more detailed clinical or family histories or underwent different clinical examination in the ER.

**Figure 1.**
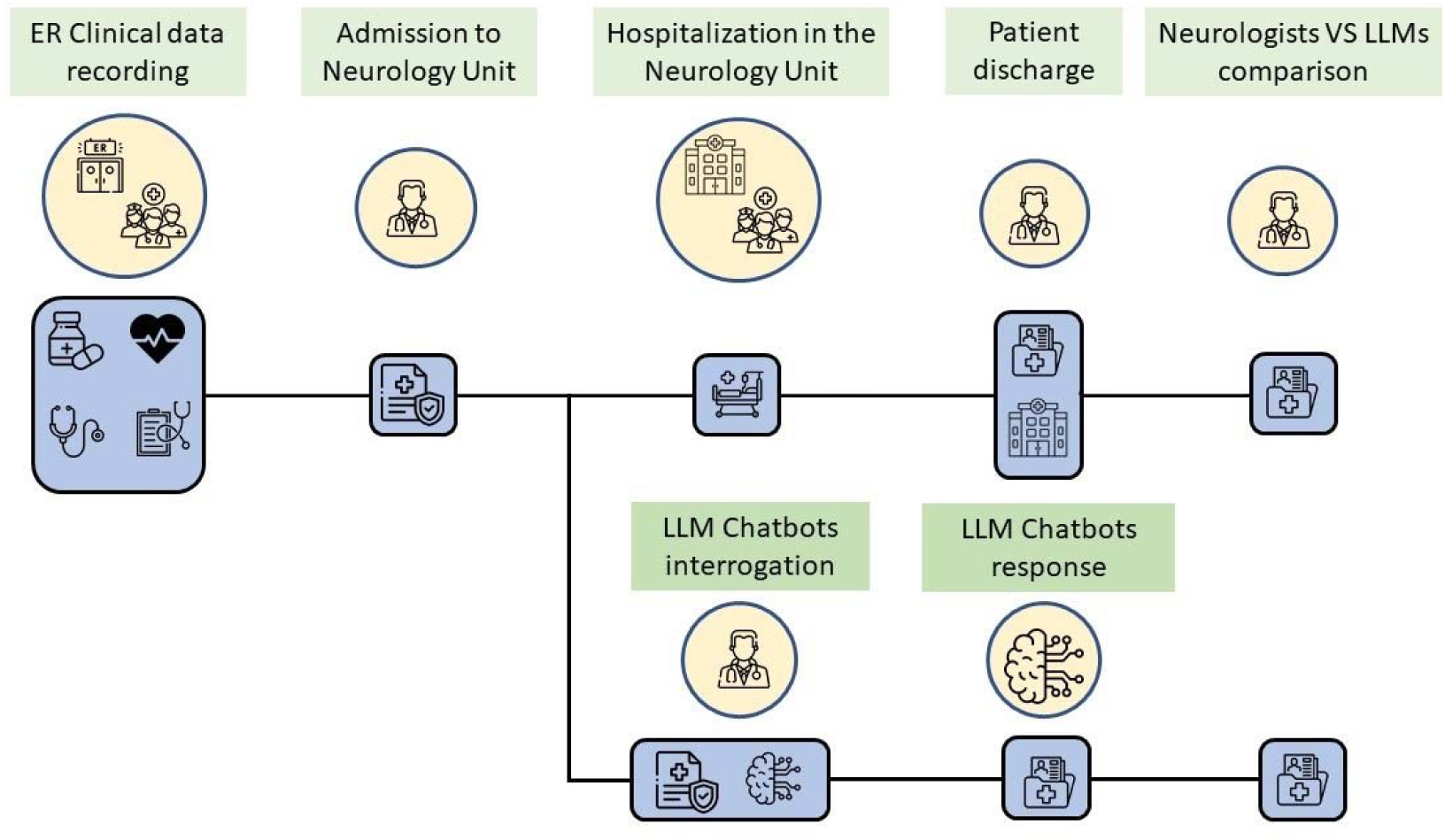
Procedure used to compare diagnosis and clinical tests assessment from neurologists and LLMs

**Figure 2.**
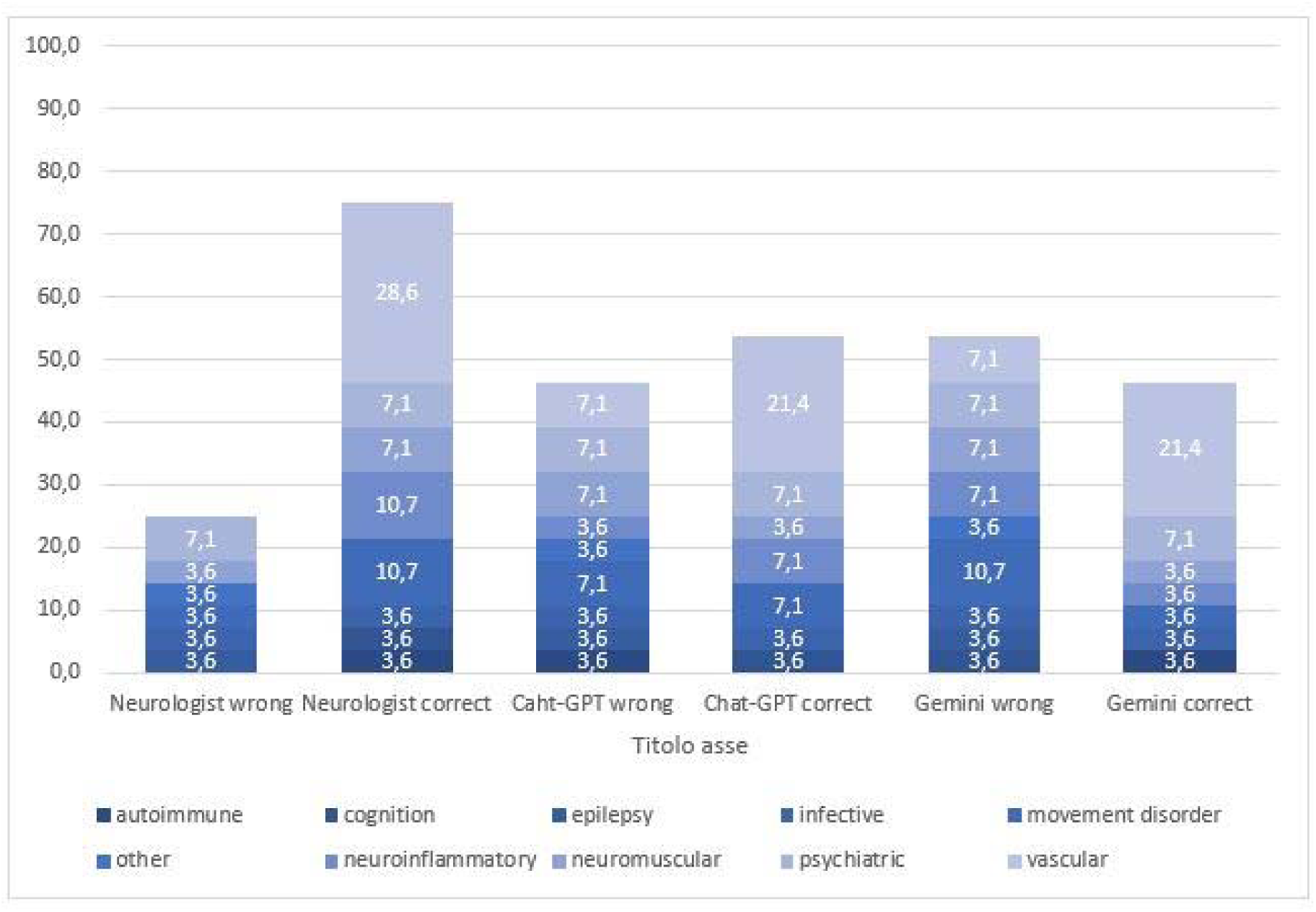
Diagnostic accuracy of Neurologists, ChatGPT-3.5, and Gemini across neurological disease categories. Results are presented as percentages of the total cases (n = 28).

#### Data presentation

Clinical cases were presented to ChatGPT-3.5 and Gemini, both in their free versions, in September 2024, to assess their reliability in providing neurological diagnoses.

The clinical cases were presented by providing the EHT from the ER, which referred patients to the neurology unit for further evaluation. This ensured that both ChatGPT-3.5 and Gemini received the same information available to neurologists at the time of admission.

Electronic health records (EHRs) were directly copied and pasted into the dialogue box of the LLMs after being fully anonymized, ensuring compliance with privacy regulations.

After the first patient case, subsequent cases were sequentially added in the same session. This methodology aimed to replicate a typical use case in which a physician interacts with the model pragmatically, focusing on obtaining responses rather than ensuring the model’s precise comprehension of the input data.

Cases were presented by an experimenter unaware of the final diagnosis made by the neurologist to both models under standardized conditions, using the following prompts:

- ChatGPT prompt: Italian version, *“Ora ti darò un caso clinico di un paziente ipotetico, dovrai indicarmi la diagnosi più probabile e una serie di esami clinici da effettuare per confermare o smentire la diagnosi, va bene?”*; English Version, *“Now I will give you a clinical case of a hypothetical patient. You will need to indicate the most likely diagnosis and suggest a series of clinical tests to confirm or rule out the diagnosis. Is that okay?”*
- Gemini prompt: Italian version, *“ora ti passerò dei casi di neurologia fittizi devi dirmi la più probabile diagnosi in base alle informazioni che ti passo e i test che faresti come approfondimento diagnostico”*; English version, *“I’m going to give you some made-up neurological cases. Your job is to tell me what you think is wrong with the patient and what tests you’d order.”*

The prompts had the specific aim to bypass core prompts designed to avoid the suggestion of potentially harmful information such in case of diagnostic and treatment indication. No other specifications on cases formatting or scheme of the response requested were given to the LLMs. The text given to LLMs was anonymized to protect patients’ privacy and not further formatted or organized, to present it as raw as possible. Both LLMs were tested on multiple cases, without additional contextual information or model-specific tuning.

The responses from ChatGPT and Gemini were recorded along with the neurologist’s suspected diagnosis at admission, the definitive discharge diagnosis, and the diagnostic tests recommended by the neurologist during the admission.

#### Evaluation

The diagnostic accuracy for the neurologists, ChatGPT, and Gemini was evaluated based on their ability to match the patient’s discharge diagnosis.

The following variables were considered for evaluation: the frequency of the cases where neurologists correctly suspected a diagnosis, confirmed as the final discharge diagnosis; the accuracy of both ChatGPT-3.5 and Gemini in suggesting the correct diagnosis; and the agreement between the neurologist, ChatGPT-3.5, and Gemini in their diagnostic indications. Additionally, it was recorder the number of times both LLMs recommended all necessary diagnostic tests, partially recommended them, or overprescribed unnecessary tests. During the whole process, the number of times in which LLMs needed more information or failed to give a valid response was recorded.

## Results

Clinical case records from 56 patients were analyzed. Twenty-eight patients (Mean age: 58.2 years; sex: 16 females) who met the inclusion criteria were selected for the study.

Both ChatGPT-3.5 and Gemini understood the prompt without requiring further specifications. ChatGPT-3.5 responded only with the request of the first clinical case, Gemini responded with an explanation of how the future responses would be organized indicating that the scheme of the response would be composed by three sections: diagnostic hypothesis, differential diagnosis and recommended diagnostic test.

After the first response ChatGPT-3.5 needed further indication in nine cases to achieve complete responses. In one case (id = 7), a clarification was requested to define whether the condition was considered primarily psychiatric or neurological. In six cases (id = 10, 12, 15, 17, 22, 28) ChatGPT-3.5 was prompted to provide both the missed diagnosis and clinical tests. In two cases (id = 6, 18) specific clinical tests were solicited to confirm or exclude a diagnosis. Conversely, Gemini generally provided an organized response framework and required explicit prompting to deliver a diagnosis only in one case (id =28).

In none of the cases ChatGPT or Gemini showed hallucination defined as a nonfactual, nonsensical, or inconsistent response to a clear given prompt ^47^.

In relation to the accuracy of the proposed diagnosis, Neurologists correctly diagnosed 75 % of cases, while ChatGPT-3.5 achieved an accuracy of 54 % and Gemini 46 %.

The diagnostic accuracy was evaluated across a range of neurological disorders, revealing notable differences in performance.

In infectious diseases, all three raters—neurologist, ChatGPT-3.5, and Gemini—demonstrated equal accuracy, correctly diagnosing 50% of the cases. For movement disorders, both neurologists and ChatGPT-3.5 correctly diagnosed 75% of the cases, while Gemini’s performance was notably lower, correctly diagnosing only 25%.

For neuromuscular disorders, neurologists, ChatGPT-3.5, and Gemini all displayed equal accuracy, correctly diagnosing 67 % of the cases, leaving 33 % of the cases misdiagnosed by each rater. In psychiatric disorders, neurologists and Gemini both achieved a 50% accuracy rate, whereas ChatGPT-3.5 underperformed, correctly diagnosing only 25% of the cases.

In vascular disorders, neurologists and Gemini both had an accuracy of 75%, while ChatGPT-3.5 performed slightly lower, with a 62 % correct diagnosis rate.

Overall, neurologists achieved the highest diagnostic accuracy across all pathologies, correctly diagnosing 75% of the cases. ChatGPT-3.5 followed with an overall accuracy of 54%, and Gemini achieved an accuracy of 46% (Fig. 3).

**Figure 3.**
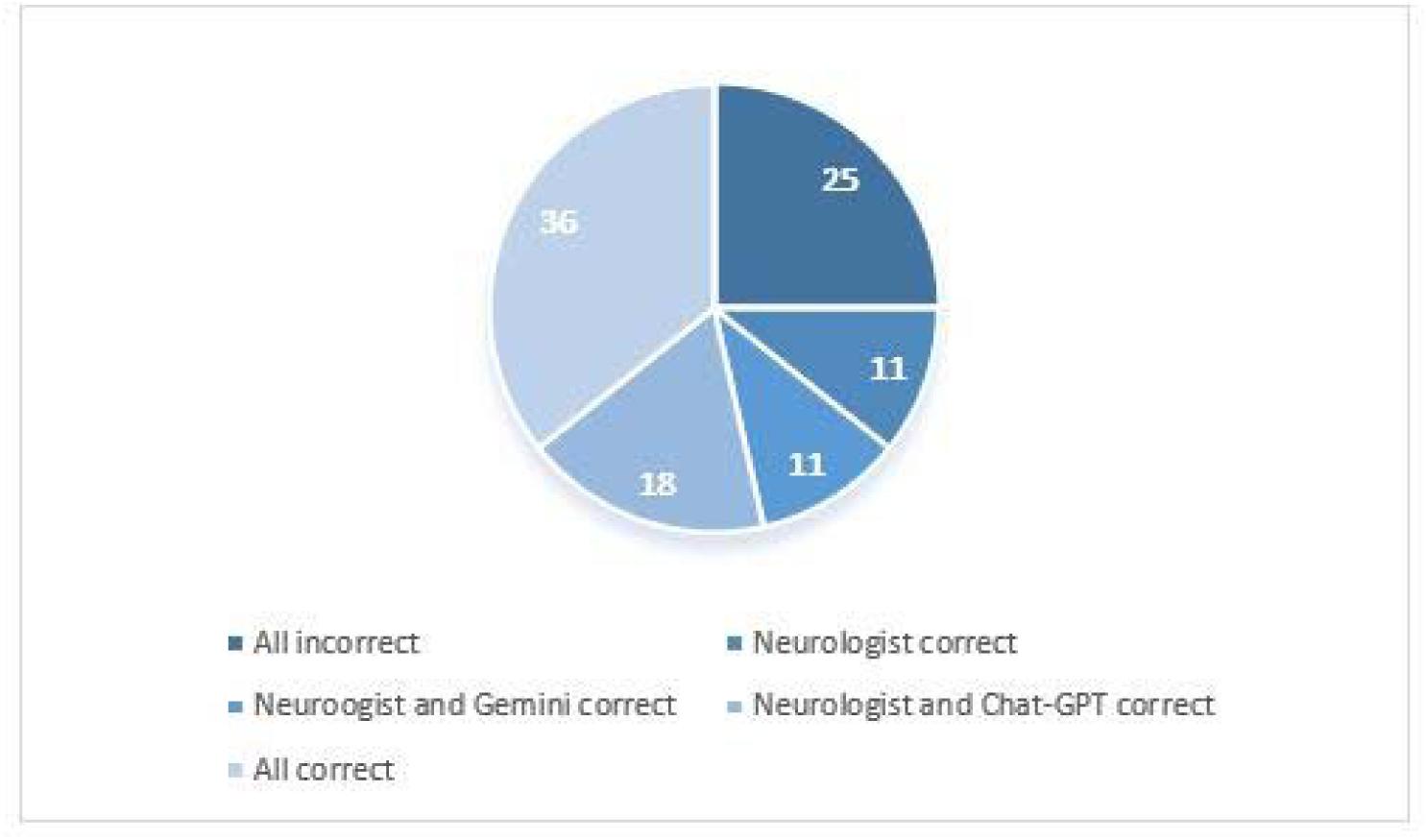
Agreement between Neurologist, ChatGPT-3.5, and Gemini in indicating correct versus incorrect diagnoses. Results are presented as percentages of total cases (n = 28).

In terms of diagnostic agreement, all three neurologists, ChatGPT-3.5, and Gemini agreed providing the correct diagnosis in 36 % of cases. In 25 % of cases, all three were incorrect. In 11% of cases only the neurologists were correct and both ChatGPT-3.5 and Gemini were incorrect. ChatGPT-3.5 only in the 11% of cases, while both the neurologist and Gemini were accurate, the percentage was 11 % of cases, and only Gemini was wrong in 18 % of cases.

In none of the cases ChatGPT-3.5 and Gemini were correct while Neurologist failed in indicating the correct diagnosis (Fig. 4).

**Figure 4.**
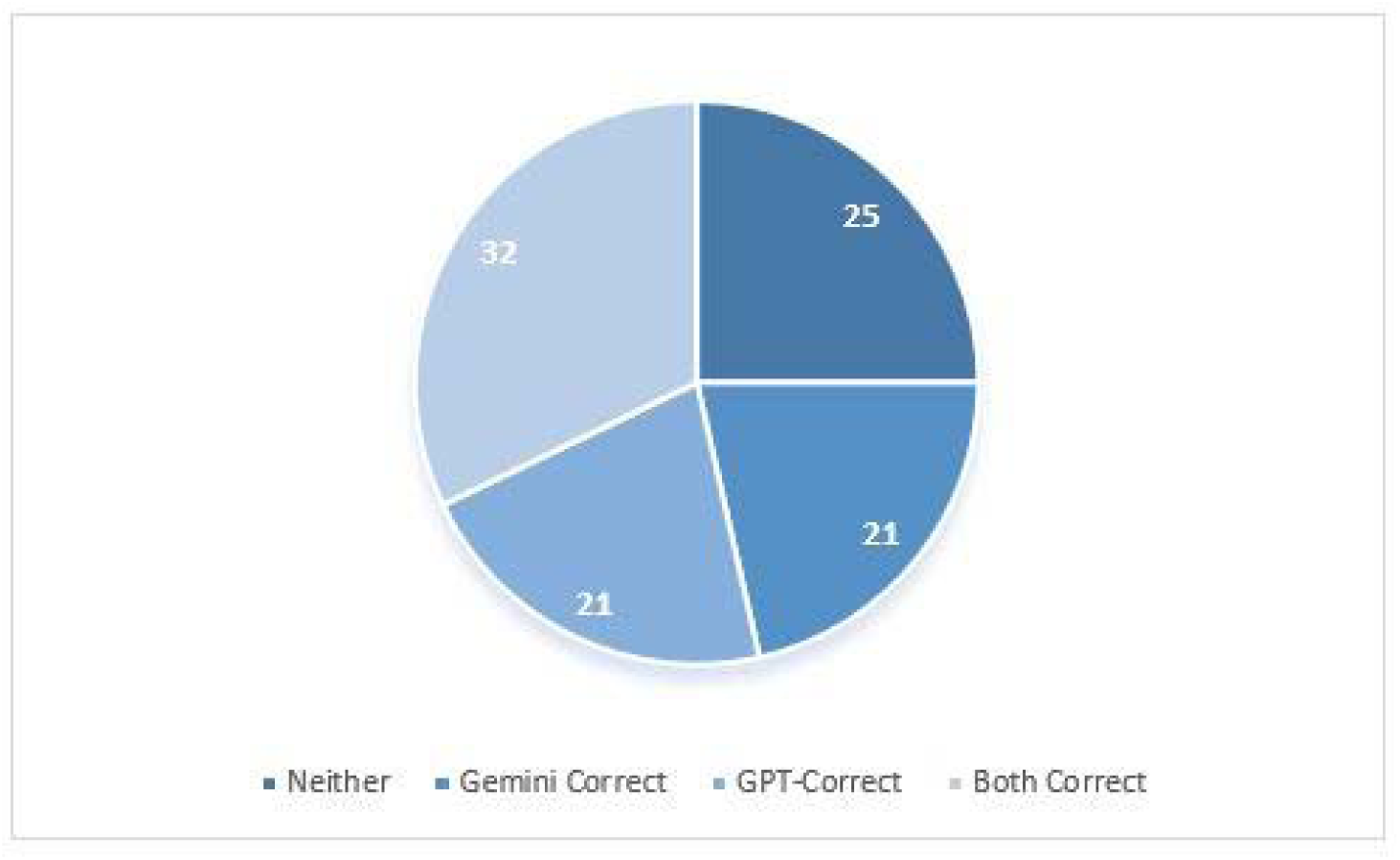
Agreement between ChatGPT-3.5, and Gemini in recommending correct versus incorrect diagnostic tests. Results are presented as percentages of total cases (n = 28).

Regarding the diagnostic test indication, both ChatGPT-3.5 and Gemini recommended the correct set of basic tests in 55 % of cases. However, there was agreement between ChatGPT-3.5 and Gemini in suggesting the correct diagnostic tests in 32 % of cases (Fig. 5).

**Figure 5.**
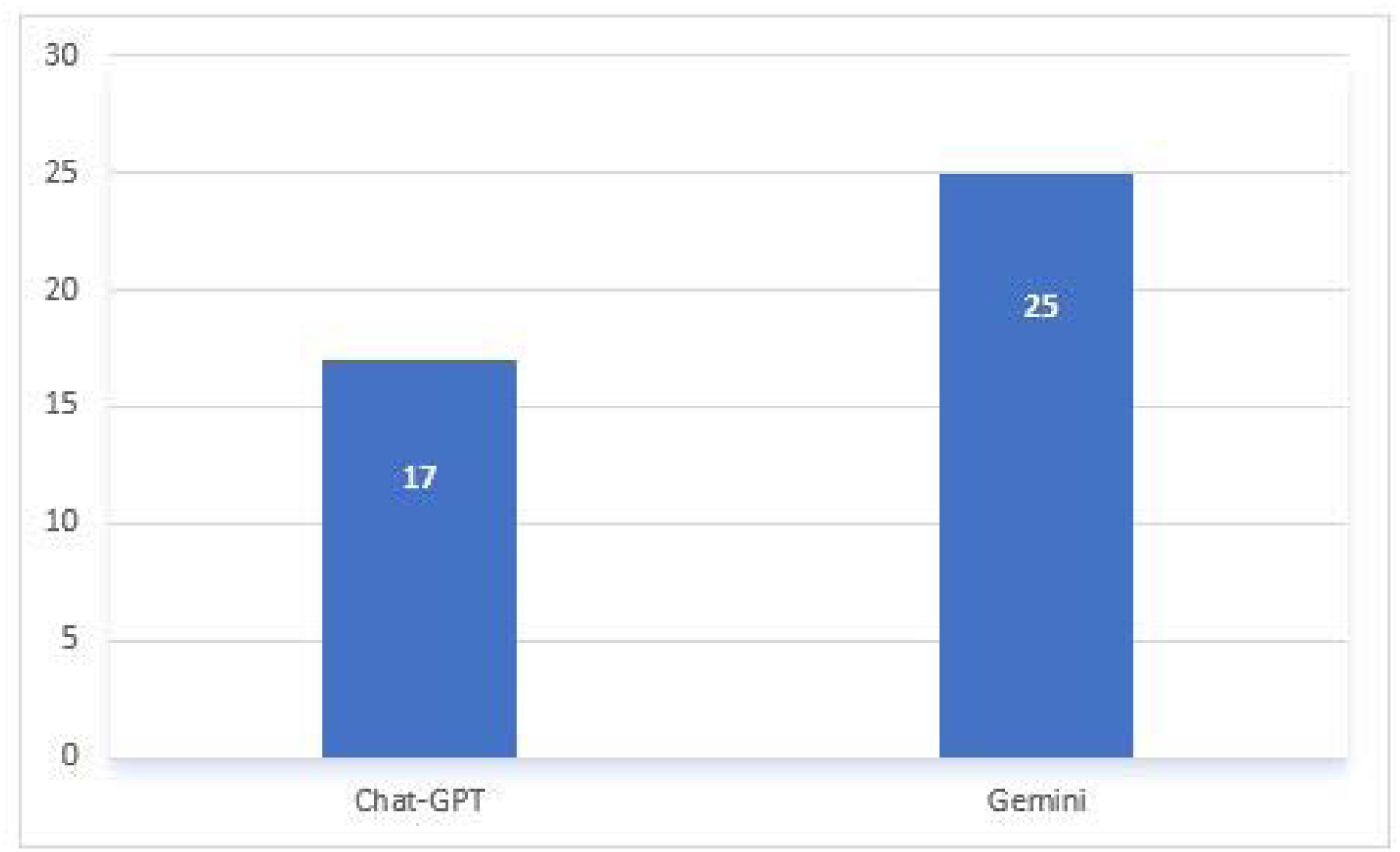
Frequency of over-prescription of diagnostic tests by ChatGPT-3.5, and Gemini. Results are presented as percentages of total cases (n = 28).

In terms of over-prescription of diagnostic tests, ChatGPT-3.5 recommended an excessive number of tests in 17 % of cases, while Gemini did so in 25% (fig. 6). In 64% of instances neither ChatGPT-3.5 or Gemini overprescribed unnecessary tests. Gemini alone over-prescribed tests in 18% of cases, ChatGPT-3.5 alone did so in 11%, and both models suggested excessive tests in 7% of cases (fig.7).

**Figure 6.**
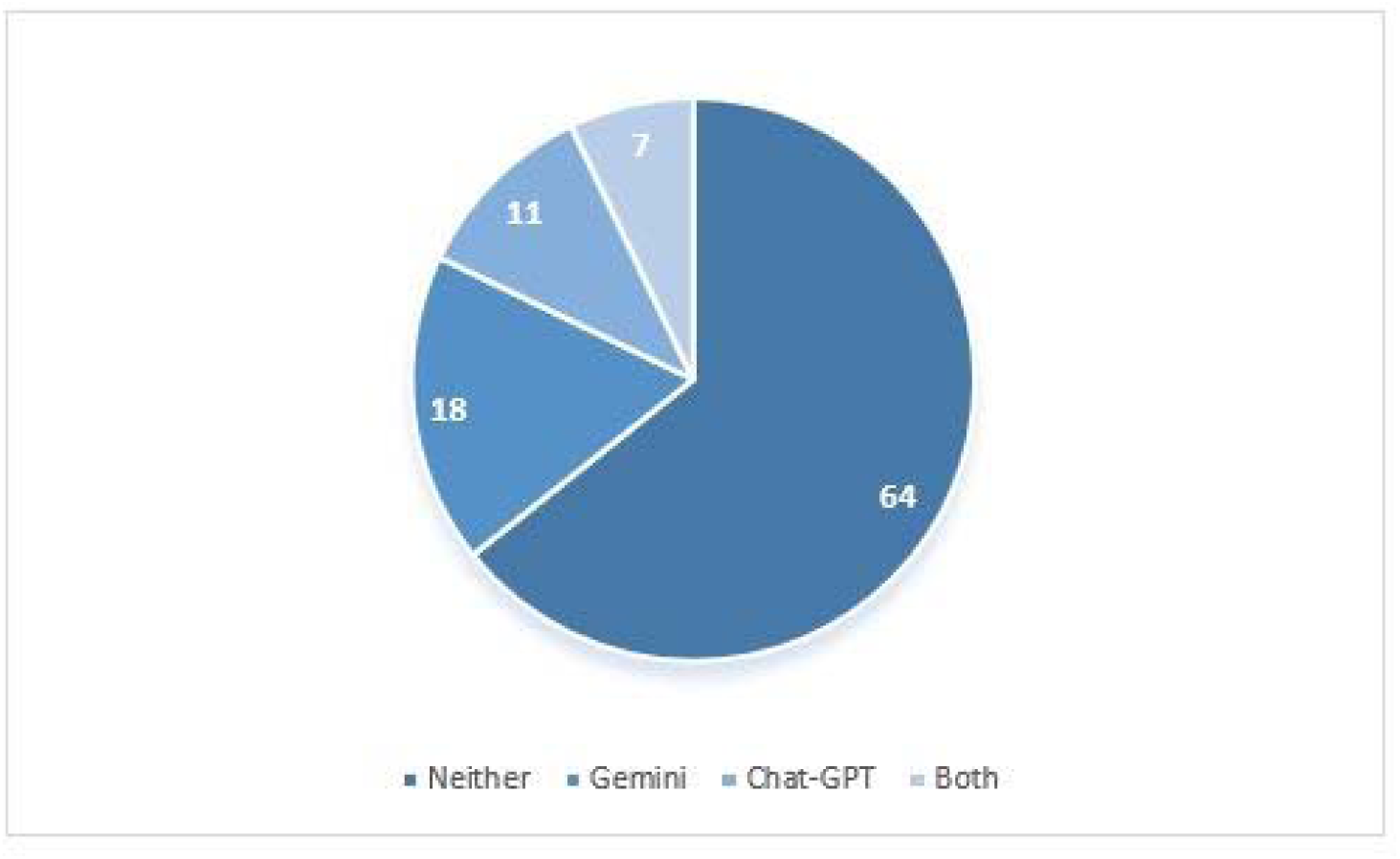
Agreement between ChatGPT-3.5, and Gemini in the over-prescription of diagnostic tests. Results are presented as percentages of total cases (n = 28).

## Discussion

This study aimed to evaluate the potential of GPT-powered LLMs (specifically ChatGPT-3.5 and Gemini). in assisting daily clinical practice within a neurological setting. A cohort of patients with diverse neurological conditions were analyzed, and the performance of the LLMs was compared to that of neurologists, thus allowing to assess the potential value of LLMs as diagnostic support in real-world scenarios. Our results showed that, in non-structured environments, in terms of diagnostic accuracy, neurologists consistently outperformed both LLMs across various neurological disorders. While all three raters achieved comparable accuracy in certain disease categories, such as infectious diseases and neuromuscular disorders, neurologists demonstrated superior performance in movement disorders, psychiatric disorders, and vascular disorders. These conditions often require nuanced clinical judgment, experience in interpreting subtle clinical signs, and the ability to integrate information ^48^. These results align with the broader literature on LLMs in specialized medical contexts, where human expertise surpasses LLMs’ performance, especially in areas requiring nuanced clinical judgment, advanced interpretative skills, and integrative diagnostic approaches ^22^. Regarding diagnostic agreement, there was substantial variability among the three raters. While all three agreed on the correct diagnosis in a significant proportion of cases, in no cases the wrong response was given by the neurologists only. Our findings align with studies showing that LLMs accuracy in clinical recommendation varies with the severity of the presentation and the complexity of the clinical picture: it performs well when initial symptoms clearly indicate a condition, but less so with more ambiguous cases ^49^. LLMs appear accurate on the surface when managing well-known, sharply defined symptoms but often fail to provide appropriate clinical management recommendations ^49^. This shortfall likely arises from the lack of deep, discipline-specific knowledge and of a clear understanding of clinical constraints, limiting their ability to navigate patient’s management effectively, beyond initial diagnosis ^49^. One crucial aspect of LLMs use regard the clinical test examination. In this scenario, both LLMs were able to recommend the appropriate basic test set in a majority of cases, however it was observed a low rate of agreement between ChatGPT-3.5 and Gemini. Over-prescription of tests was observed for both models, especially for Gemini. This tendency underscores a limitation shared by LLMs in clinical applications which is the inability to apply cost-benefit reasoning in patient care.

While emphasizing the promise of LLMs in clinical practice, it should be noted their limitations in handling complex diagnosis and in recommendation of further diagnostic assessment.

LLMs can support clinicians in structured assessment contexts, but they are still limited in their ability to account for the subtle and often ambiguous nature of real-world clinical decision-making^48^.

It should be considered that LLMs rely on training data that could be biased thus influencing diagnostic suggestions, potentially resulting in skewed or less reliable outputs ^14^. In addition, other studies observed that LLMs have limitations reflecting data-driven biases, particularly in domains requiring high sensitivity to demographic or clinical subcategories ^50^.

## Conclusions

In conclusion, while our findings indicate that LLMs hold potential as supportive tools in clinical neurology, they cannot yet replace the depth of human expertise and should be used with strict human supervision ^48^.

While both the models analyzed (ChatGPT-3.5 and Gemini) demonstrated a remarkable ability to comprehend complex clinical scenarios and generate relevant responses, significant differences emerged in their performance and responses style. ChatGPT-3.5, while capable of providing accurate diagnoses and test recommendations, often required additional prompting to deliver comprehensive and objective answers. In contrast, Gemini exhibited a more structured approach, providing a clear framework for its responses and requiring explicit prompting for specific diagnostic conclusions. These factors could be of fundamental importance in the evaluation of the usability of the systems and should be kept into consideration when implementing LLMs in the real practice ^51^. The ability of these models to adopt a human-like interaction pattern can be advantageous in human-machine interaction. However, this same feature also increases the risk of misunderstandings due to natural language, unlike typical clinical decision support systems, which rely on standardized and unambiguous communication protocols. The use of LLMs requires careful management to ensure consistency and prevent misinterpretation of the information they provide. New studies should focus on how models are utilized by clinicians within a human-in-the-loop framework, testing how experts use LLMs. For example, a study by Goh and colleagues ^52^ showed that while LLMs alone performed better than clinicians using conventional resources, integrating LLMs with expert oversight could enhance diagnostic accuracy and efficiency indicating the potential for significant improvements in clinical practice when AI tools are effectively integrated with human expertise.

Future work should focus on refining LLMs’ algorithms to better interpret clinical data and test the reliability of responses using large datasets of real-world clinical cases, creating models that complement human expertise. By addressing these areas, LLMs may progressively evolve into valuable tools for enhancing diagnostic precision and supporting clinical decision-making. LLMs are particularly useful in time-sensitive situations and as educational tools for junior clinicians. However, when faced with nuanced or ambiguous cases that require integration of subtle clinical cues, LLMs often fail. Their limitations in handling complexity, making cost-effective recommendations, and avoiding over-prescription of diagnostic tests highlight the current gap between AI performance and the demands of real-world clinical practice: in clinical settings, the unpredictability and high-pressure nature of day-to-day operations require models that can handle dynamic challenges and operate effectively under less-than-ideal conditions. Therefore, it is crucial to bridge the gap between laboratory testing and real-world application to ensure LLMs are equipped to meet the demands of clinical practice.

## Data Availability

All data produced in the present work are contained in the manuscript

## Notes

### Competing Interest Statement

The authors have declared no competing interest.

### Funding Statement

This study did not receive any funding

### Author Declarations

The present study qualifies as a non-interventional observational study and does not constitute a clinical trial. The study has been approved by the IRB of the University of Milan with the approval number 123/24, All. 3 CE 10/12/24. Patients provided consent for the processing of their personal data at the time of hospital admission under protocol ast_daz_502_ed00, in accordance with privacy regulations (D.Lgs. 101/2018, implementing EU GDPR 2016/679). This consent permits research on clinical data. The data have been anonymized prior to processing, ensuring that they cannot be traced back to any specific patient.

